# Gabapentin for Postoperative Pain Management in Lumbar Fusion Surgery

**DOI:** 10.1101/2022.09.20.22280097

**Authors:** Eli A Perez, Emanuel Ray, Brian J Park, Colin J Gold, Ryan M Carnahan, Matthew Banks, Robert D. Sanders, Catherine R Olinger, Rashmi N Mueller, Royce W Woodroffe

## Abstract

**Study Design:** Retrospective cohort study

**Objectives:** To investigate the effect of perioperative gabapentin administration on postoperative pain and opioid use in elderly patients after elective posterior lumbar fusion.

**Methods:** This was a single-center study of patients over the age of 65 who underwent elective posterior lumbar fusion. The cohort was stratified into two groups based on perioperative gabapentin administration defined as use both in the immediate preoperative period and postoperatively during hospitalization. Mean daily visual analogue scale (VAS) pain scores and mean daily morphine equivalent dosage (MED) during the postoperative hospital stay were compared between these groups using traditional regression and inverse probability weighting.

**Results:** There were 350 patients who received gabapentin perioperatively and 78 patients who did not receive gabapentin. Multivariate regression did not demonstrate a difference in daily MED (*ß=-6.6 mg, p=0.816*) or daily pain scores (*ß=0.04, p=0*.257) averaged over the first seven postoperative days. Sub-analysis stratifying by each individual postoperative day also did not demonstrate a difference in MED or pain scores.

**Conclusion:** This study did not find perioperative gabapentin administration to be beneficial in reducing postoperative VAS pain scores or opioid use in the acute postoperative period for elderly patients undergoing posterior lumbar fusion. These findings are consistent with recent literature suggesting gabapentin is of minimal benefit in postoperative multimodal analgesia regimens.

**Level of Evidence:** 3

## INTRODUCTION

Opioid medications remain the cornerstone of postoperative pain management despite decades of research into the pathophysiology of pain and alternative therapeutics.^1^ Although beneficial in many aspects, opioids have several adverse effects in the postoperative period including nausea, ileus, and delirium.^2-4^ Postoperative opioid use is also associated with long-term dependence with up to 27% of opioid-dependent patients having their first exposure to opioids during the perioperative period.^4,5^ Owing to these risks, the use of opioids for postoperative pain has been under increased scrutiny in recent years.^6^

Concerns regarding pain management and other aspects of perioperative care have led to the development of enhanced recovery after surgery (ERAS) protocols that aim to improve perioperative outcomes through the use of evidence-based practices.^7-10^ Many ERAS protocols implement multi-modal analgesia (MMA) regimens with the goal of lowering opioid usage, thus avoiding opioid-related adverse events while maintaining adequate pain control.^11^ MMA regimens combine several pain management strategies including utilizing alternative pain relief medications such as gabapentinoids, muscle relaxants, and acetaminophen.^12,13^

Gabapentinoids have received a significant amount of attention as an alternative to opioids. Gabapentin, the oldest drug in its class, is the most well-studied for its role in pain management. Although questions remain regarding the cellular mechanisms, current evidence suggests that gabapentinoids inhibit neurotransmitter signaling which ultimately blocks the peripheral and central sensitization of the nervous system to nociceptive signaling thereby alleviating pain.^14-16^ These mechanisms were found to play a prominent role in the pathogenesis of chronic pain where gabapentin has been shown to be effective.^17,18^ Studies later suggested that peripheral and central sensitization also occurred with acute pain which provided a theoretical basis for gabapentin’s use in postoperative pain management.^16,19,20^ Despite mechanistic support for its use in acute pain, clinical studies investigating the effect of gabapentin in perioperative pain control and opioid reduction have been mixed. Early studies showed it successfully decreases opioid usage in the acute postoperative period, especially in the first 24 to 48 hours.^12,21-24^ However, more recent evidence suggests gabapentin has minimal impact on post-operative pain scores or opioid usage and may, in fact, increase the risk of adverse effects.^25,26^

Much of the literature regarding gabapentin’s benefits in treating postoperative pain have focused on patients undergoing general surgery. Few studies have focused on the spine surgery population, and of those that have been performed, evidence suggests that gabapentin is beneficial.^27-29^ However, given this conflicts with recent surgical literature, there is a need for further investigation.^26^ The purpose of the current study is to investigate the efficacy of gabapentin in pain management during the postoperative hospital course in patients who undergo posterior lumbar fusion.

## PATIENTS AND METHODS

### Study Design

Following institutional review board approval, a retrospective analysis of our institution’s electronic medical record was performed for patients who underwent posterior lumbar fusion performed by neurosurgeons and orthopedic surgeons between January 1, 2017 and February 1, 2021. This study included patients with degenerative spine disease who were over 65 years of age at the time of surgery and had undergone elective surgery. Patients who did not have postoperative pain scores recorded or who underwent emergent surgery were excluded from this study.

The MMA protocol used for spine surgery patients at our institution is predominantly standardized with some variations based on the attending surgeon’s preference. Generally, patients receive 600 mg of gabapentin with or without a 975 mg acetaminophen loading dose approximately 30 minutes prior to surgery. This is followed by postoperative administration of acetaminophen, gabapentin, and/or a muscle relaxant (baclofen or cyclobenzaprine) in addition to standard opioid medications. These postoperative medications are administered as needed to achieve adequate pain control.

The study cohort was divided into two groups based on whether they received gabapentin as part of these MMA protocols. The first group received the 600 mg dose of gabapentin preoperatively and continued to receive gabapentin postoperatively as part of their MMA regimen. The second group received the same MMA regimen but did not receive either preoperative or postoperative gabapentin. A subset of patients in the initial cohort had inconsistent gabapentin administration in the hospital (i.e., they received gabapentin preoperatively but did not receive it postoperatively) and were excluded.

### Data Collection and Outcome Measures

Basic demographic factors at the time of surgery were collected from the medical records including age, gender, history of diabetes, history of hypertension, and smoking status. In addition, preoperative pain medication (opioid, gabapentinoid, muscle relaxant, acetaminophen) and psychiatric medication (anti-depressants, benzodiazepines, non-benzodiazepines) usage was also recorded.

Data collected from the perioperative hospital course included the time under anesthesia, length of surgery, number of interbody spacers placed, vertebral levels fused, American Society of Anesthesiologists (ASA) status, daily postoperative gabapentin dosage, postoperative muscle relaxant administration, use of patient-controlled analgesia (PCA), acetaminophen administration in day of surgical admission (DOSA), and daily oral and parenteral opioid use. Opioid dosing was converted to morphine equivalent dosage (MED) for analysis. A list of conversion factors is provided in Appendix A.

The primary outcomes were average daily MED and average daily pain scores for up to seven days postoperatively. Since length of stay impacted the available data for opioid use and pain, an alternative outcome that limited the data to the first three postoperative days was also explored. Postoperative day 1 was defined as starting at midnight the day after surgery. Postoperative opioid use was determined based on recorded doses of parenteral and oral opioid medications. Pain scores were recorded by surgical ward nurses. Secondary outcomes included length of stay, disposition at discharge, surgical ICU length of stay, infection rates, all-cause reoperation rates, and postoperative delirium.

### Statistical Analysis

Descriptive statistics were used to compare study groups. Categorical variables were summarized with frequencies and compared using Pearson’s chi-square test or a Fisher’s exact test for frequencies less than 5. Continuous variables were summarized with means and standard deviations and analyzed using unpaired, two-tailed Welch’s t-test. Traditional multivariate analysis was used to compare the relationship between gabapentin use and study outcomes while controlling for confounders. The selection of predictor variables for each model was based on a univariate significance cutoff of *p <0.2* and prior literature. Variables demonstrating multicollinearity were excluded from these models. Continuous outcomes were analyzed using linear regression, while logistic regression was used for categorical outcomes.

An additional exploratory analysis investigating mean MED and mean VAS pain scores over the first three postoperative days was performed utilizing doubly robust inverse probability weighted (IPW) models with stabilized average treatment effect weights to reduce the risk of bias from study design. Generalized estimating equations with empirical standard error estimates were used for doubly robust models to account for weighting. Patients with propensity scores outside the region of common support were excluded from this analysis.

Statistical analysis was performed using R Statistical Software, Version 4.0.4 (Vienna, Austria: R Foundation) and SAS/STAT software, version 9.4 of the SAS system for Windows (Copyright 2016, SAS Institute, Inc., Cary, NC, USA). Statistical significance was established at *p <0.05*.

## RESULTS

### Cohort Characteristics

Initial screening found 702 patients with 428 meeting the inclusion and exclusion criteria. The gabapentin group included 350 patients while the no gabapentin group included 78 patients. A comparison of baseline demographics for these groups is provided in Table 1. Univariate analysis found no statistically significant difference between gender distribution and mean age. Both groups had a similar number of fusion levels; however, the no gabapentin group had significantly longer time under anesthesia (*p* = 0.009), longer length of surgery (*p* = 0.007), and greater number of interbodies placed (*p* = 0.018). The gabapentin group was more likely to receive acetaminophen in day of surgical admission (*p* < 0.001) as well as PCA opioids (*p* < 0.001), while postoperative muscle relaxant use was similar in both groups. A comparison of preoperative outpatient medication usage (Table 2) found the gabapentin group to have a higher proportion of preoperative gabapentinoid use (*p* < 0.001). Both groups otherwise had similar prescription rates for antidepressants and opioids.

**Table 1.** Cohort Demographics Stratified by Gabapentin Usage.

|  | No Gabapentin<br>(n=78) | Gabapentin<br>(n=350) | P-value<sup>a</sup> |
| --- | --- | --- | --- |
| Age, mean (SD) | 73.2 (5.8) | 72.0 (5.3) | 0.104 |
| Diabetes, n (%) | 15 (19.2%) | 83 (23.7%) | 0.394 |
| Depression History, n (%) | 4 (1.1%) | 15 (4.3%) | 0.761 |
| Hypertension, n (%) | 43 (55.1%) | 199 (56.9%) | 0.781 |
| Gender |  |  |  |
| Male, n (%) | 39 (50%) | 166 (47.4%) | 0.681 |
| Female, n (%) | 39 (50%) | 184 (52.6%) |  |
| Smoking History, n (%) | 28 (35.9%) | 165 (47.1%) | 0.071 |
| Smokeless Tobacco Use, n (%) | 2 (2.6%) | 26 (7.4%) | 0.135 |
| Time in Surgery in minutes,<br>mean (SD) | 255.9 (105.5) | 221.7 (92.7) | **0.007** |
| Time Under Anesthesia<br>minutes, mean (SD) | 352.0 (80.4) | 324.4 (76.0) | **0.009** |
| ASA, n (%) |  |  | 0.348 |
| Grade 1 | 0 | 2 (0.6%) |  |
| Grade 2 | 29 (37.2%) | 150 (42.9%) |  |
| Grade 3 | 46 (59.0%) | 193 (55.1%) |  |
| Grade 4 | 3 (3.8%) | 5 (1.4%) |  |
| Grade 5 | 0 | 0 |  |
| Number of Levels, mean (SD) | 3.9 (2.1) | 4.0 (2.5) | 0.828 |
| Number of interbodies, mean<br>(SD) | 0.962 (1.0) | 0.657 (0.950) | **0.018** |
| Acetaminophen in DOSA, n<br>(%) | 27 (7.7%) | 334 (95.4%) | **< 0.001** |
| Postoperative Muscle<br>Relaxant, n (%) | 53 (67.9%) | 231 (66.0%) | 0.742 |
| Postoperative PCA, n (%) | 21 (26.9%) | 271 (77.4%) | **< 0.001** |
| <sup>a</sup>Unadjusted P-values reported |  |  |  |
| <sup>b</sup>Abbreviations: American Society of Anesthesiologists (ASA), Day of Surgical Admission (DOSA), Patient-Controlled Analgesia (PCA) |  |  |  |

**Table 2.** Preoperative Outpatient Medications.

|  | **No Gabapentin**<br>**(n=78)** | **Gabapentin**<br>**(n=350)** | **P-value<sup>a</sup>** |
| --- | --- | --- | --- |
| **Antidepressant<sup>b</sup>, n (%)** | 27 (34.6%) | 144 (41.1%) | 0.287 |
| **Atypical, n (%)** | 7 (9.0%) | 53 (15.1%) | 0.156 |
| **TCA, n (%)** | 4 (5.1%) | 19 (5.4%) | 1.000 |
| **SSRI, n (%)** | 10 (12.8%) | 52 (14.9%) | 0.644 |
| **SNRI, n (%)** | 17 (21.8%) | 61 (17.4%) | 0.366 |
| **MAOI, n (%)** | 0 | 0 | 1.000 |
| **Opioids, n (%)** | 43 (55.1%) | 171 (48.9%) | 0.317 |
| **Gabapentinoid, n (%)** | 18 (23.1%) | 191 (54.6%) | **< 0.001** |
| **Pregablin, n (%)** | 11 (14.1%) | 7 (2.0%) | **< 0.001** |
| **Gabapentin, n (%)** | 9 (11.5%) | 187 (53.4%) | **< 0.001** |
| **Muscle Relaxant, n (%)** | 19 (24.4%) | 74 (21.1%) | 0.533 |
| **Antihistamine, n (%)** | 9 (11.5%) | 33 (9.4%) | 0.571 |
| **Barbiturate, n (%)** | 0 | 3 (0.9%) | 1.000 |
| **Benzodiazepine-like, n (%)** | 3 (3.8%) | 18 (5.1%) | 0.779 |
| **Benzodiazepines, n (%)** | 9 (11.5%) | 44 (12.6%) | 0.802 |
| **Acetaminophen, n (%)** | 45 (57.7%) | 213 (60.9%) | 0.606 |
| <sup>a</sup>Unadjusted P-values reported |  |  |  |
| <sup>b</sup>A small portion of patients were on multiple antidepressants or gabapentinoids resulting in a discrepancy in total values presented |  |  |  |

### Primary Outcomes

Opioid use and VAS pain scores were compared between study groups using multivariate regression (Tables 3 and 4). The results of the regression models for the primary outcomes of overall mean inpatient MED and VAS pain score are provided in Table 4. The variables controlled for in these regression models are also provided in Table 4. Additional sub-analysis comparing individual postoperative days was also performed (Table 3). Patients were excluded from the individual postoperative day sub-analysis regression models after their postoperative day of discharge.

**Table 3.** Opioid Use (in MED) and Post-op Pain VAS Scores by Postoperative Day (POD)

|  | Group | POD1 | POD2 | POD3 | POD4 | POD5 | POD6 | POD7 | Overall Average |
| --- | --- | --- | --- | --- | --- | --- | --- | --- | --- |
| **Pain (Mean, SD)** | **No Gabapentin** | 4.3<br>(1.5) | 4.2<br>(1.8) | 3.7<br>(1.8) | 4.0<br>(1.7) | 4.1<br>(1.9) | 3.6<br>(2.1) | 3.4<br>(1.9) | 4.0<br>(1.3) |
|  | **Gabapentin** | 4.5<br>(2.0) | 4.4<br>(1.9) | 4.2<br>(2.0) | 4.3<br>(2.1) | 4.3<br>(2.0) | 4.4<br>(2.2) | 4.7<br>(2.6) | 4.2<br>(1.7) |
|  | **P-value<sup>a</sup>** | 0.735 | 0.986 | 0.593 | 0.614 | 0.848 | 0.281 | 0.168 | 0.816 |
| **Morphine Equivalent Dose (Mean, SD)** | **No Gabapentin** | 77.0<br>(67.8) | 54.1<br>(36.4) | 39.3<br>(31.0) | 39.1<br>(38.8) | 33.2<br>(28.9) | 27.9<br>(26.1) | 22.9<br>(29.9) | 49.0<br>(30.5) |
|  | **Gabapentin** | 85.4<br>(74.4) | 51.7<br>(59.8) | 43.0<br>(72.1) | 43.0<br>(70.2) | 44.9<br>(73.8) | 43.7<br>(71.1) | 50.3<br>(75.5) | 52.0<br>(52.8) |
|  | **P-value<sup>a</sup>** | 0.149 | 0.288 | 0.802 | 0.385 | 0.928 | 0.934 | 0.828 | 0.257 |

<sup>a</sup>P-values reported for multivariate regression
<sup>a</sup>P-values reported for multivariate regression

**Table 4.**
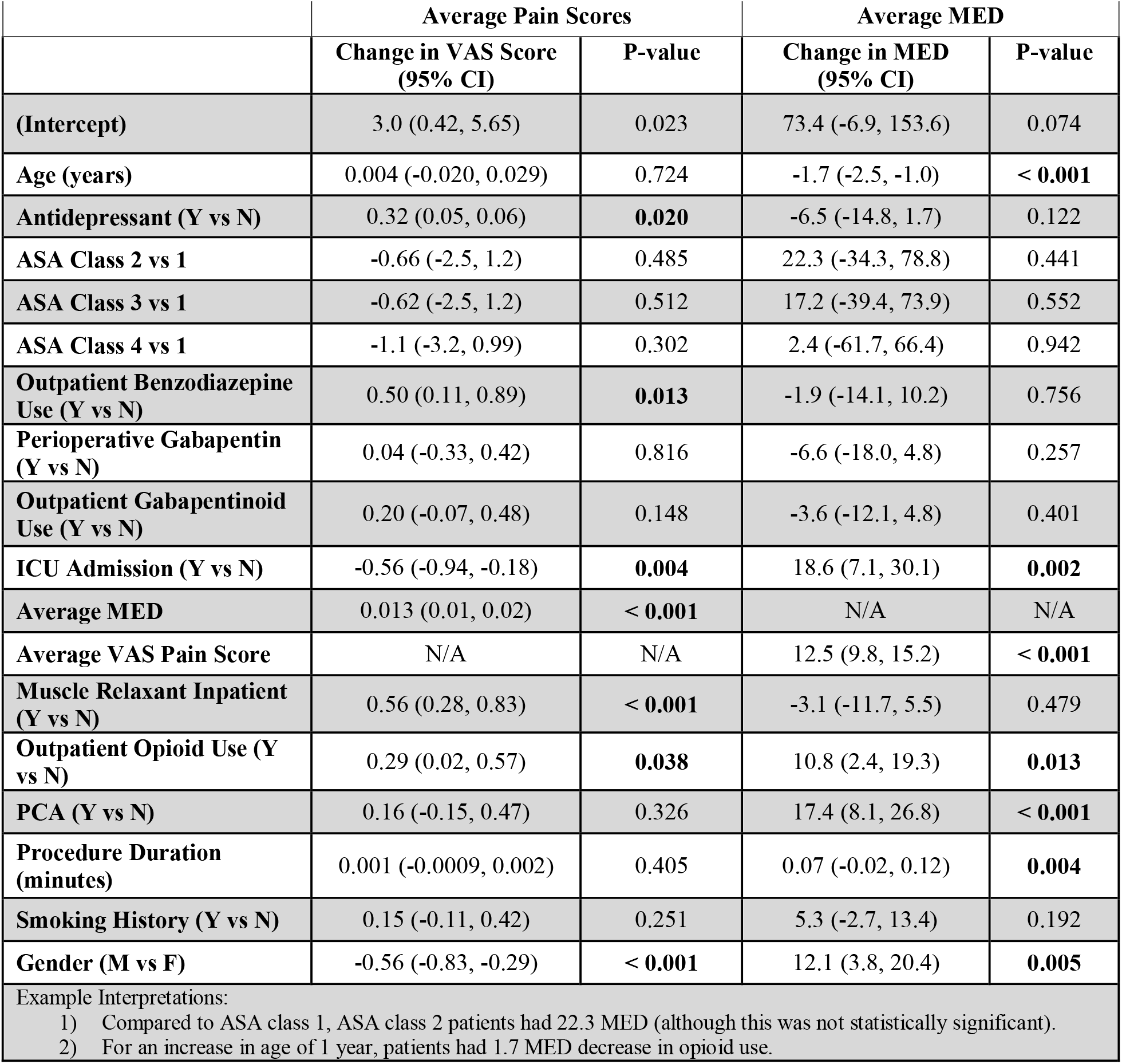
Linear Regression Models for Average VAS Pain Scores and Average MED.

|  | Average Pain Scores |  | Average MED |  |
| --- | --- | --- | --- | --- |
|  | Change in VAS Score<br>(95% CI) | P-value | Change in MED<br>(95% CI) | P-value |
| **(Intercept)** | 3.0 (0.42, 5.65) | 0.023 | 73.4 (-6.9, 153.6) | 0.074 |
| **Age (years)** | 0.004 (-0.020, 0.029) | 0.724 | -1.7 (-2.5, -1.0) | **< 0.001** |
| **Antidepressant (Y vs N)** | 0.32 (0.05, 0.06) | **0.020** | -6.5 (-14.8, 1.7) | 0.122 |
| **ASA Class 2 vs 1** | -0.66 (-2.5, 1.2) | 0.485 | 22.3 (-34.3, 78.8) | 0.441 |
| **ASA Class 3 vs 1** | -0.62 (-2.5, 1.2) | 0.512 | 17.2 (-39.4, 73.9) | 0.552 |
| **ASA Class 4 vs 1** | -1.1 (-3.2, 0.99) | 0.302 | 2.4 (-61.7, 66.4) | 0.942 |
| **Outpatient Benzodiazepine Use (Y vs N)** | 0.50 (0.11, 0.89) | **0.013** | -1.9 (-14.1, 10.2) | 0.756 |
| **Perioperative Gabapentin (Y vs N)** | 0.04 (-0.33, 0.42) | 0.816 | -6.6 (-18.0, 4.8) | 0.257 |
| **Outpatient Gabapentinoid Use (Y vs N)** | 0.20 (-0.07, 0.48) | 0.148 | -3.6 (-12.1, 4.8) | 0.401 |
| **ICU Admission (Y vs N)** | -0.56 (-0.94, -0.18) | **0.004** | 18.6 (7.1, 30.1) | **0.002** |
| **Average MED** | 0.013 (0.01, 0.02) | **< 0.001** | N/A | N/A |
| **Average VAS Pain Score** | N/A | N/A | 12.5 (9.8, 15.2) | **< 0.001** |
| **Muscle Relaxant Inpatient (Y vs N)** | 0.56 (0.28, 0.83) | **< 0.001** | -3.1 (-11.7, 5.5) | 0.479 |
| **Outpatient Opioid Use (Y vs N)** | 0.29 (0.02, 0.57) | **0.038** | 10.8 (2.4, 19.3) | **0.013** |
| **PCA (Y vs N)** | 0.16 (-0.15, 0.47) | 0.326 | 17.4 (8.1, 26.8) | **< 0.001** |
| **Procedure Duration (minutes)** | 0.001 (-0.0009, 0.002) | 0.405 | 0.07 (-0.02, 0.12) | **0.004** |
| **Smoking History (Y vs N)** | 0.15 (-0.11, 0.42) | 0.251 | 5.3 (-2.7, 13.4) | 0.192 |
| **Gender (M vs F)** | -0.56 (-0.83, -0.29) | **< 0.001** | 12.1 (3.8, 20.4) | **0.005** |
| Example Interpretations: |  |  |  |  |
| 1) Compared to ASA class 1, ASA class 2 patients had 22.3 MED (although this was not statistically significant). |  |  |  |  |
| 2) For an increase in age of 1 year, patients had 1.7 MED decrease in opioid use. |  |  |  |  |

The traditional regression model investigating the impact of perioperative gabapentin on the overall mean daily postoperative pain was well fit to the data (*R*^*2*^*= 0.3363, p* < 0.001) and did not show a difference in pain scores between the two groups (*ß=0.04, p= 0.816*). The second model comparing overall mean daily postoperative MED between the two study groups was also well fit to the data (*R*^*2*^*= 0.3454, p* < 0.001) and did not demonstrate a difference in opioid usage (*ß=-6.6, p=0.257*). The sub-analysis for individual postoperative days showed similar pain scores between the two groups in the immediate postoperative period with slightly higher pain scores on postoperative days 6 and 7, although this did not reach statistical significance. The gabapentin group also tended to have higher opioid usage on any individual postoperative day; however, this was also not statistically significant.

Exploratory analysis utilizing doubly robust IPW models was performed to compare mean MED and mean VAS pain over the first three postoperative days. The IPW VAS pain score model used 387 patients while the IPW MED model used 398 patients following trimming of outliers. The IPW VAS pain score model included acetaminophen administration in DOSA, preoperative benzodiazepine use, preoperative serotonin-norepinephrine reuptake inhibitor (SNRI) use, preoperative opioid use, hypertension history, gender, smoking history, muscle relaxant administration starting on postoperative day 1, and log mean MED. The IPW MED model was analyzed by converting mean MED to logarithm form then controlling for PCA use, preoperative atypical antidepressant use, preoperative SNRI use, preoperative muscle relaxant use, preoperative gabapentinoid use, preoperative opioid use, gender, muscle relaxant administration starting on postoperative day 1, and acetaminophen administration in DOSA. These models did not find a difference between study groups for VAS pain (*ß=0.04, p=0.8406*) or MED (*ß=1.61, p=0.460*).

### Secondary Outcomes

Multivariate regression models were developed using linear regression or logistic regression as applicable to compare the impact of perioperative gabapentin on secondary outcomes (Table 5). There was no difference in length of stay (*p=0.715)*, ICU length of stay (*p=0.715*), ICU admission rates (1.09 OR, *p=0.715*), or rates of delirium (1.31 OR, *p=0.715)* between study groups. However, the group that received gabapentin was more likely to be discharged to a skilled care facility or acute rehabilitation (2.1 OR, *p=0.036*). Due to the small sample size of patients who developed delirium or were admitted to the ICU, the mean number of days with delirium and total ICU admission time were compared using univariate analysis which did not find a difference between study groups.

**Table 5.**
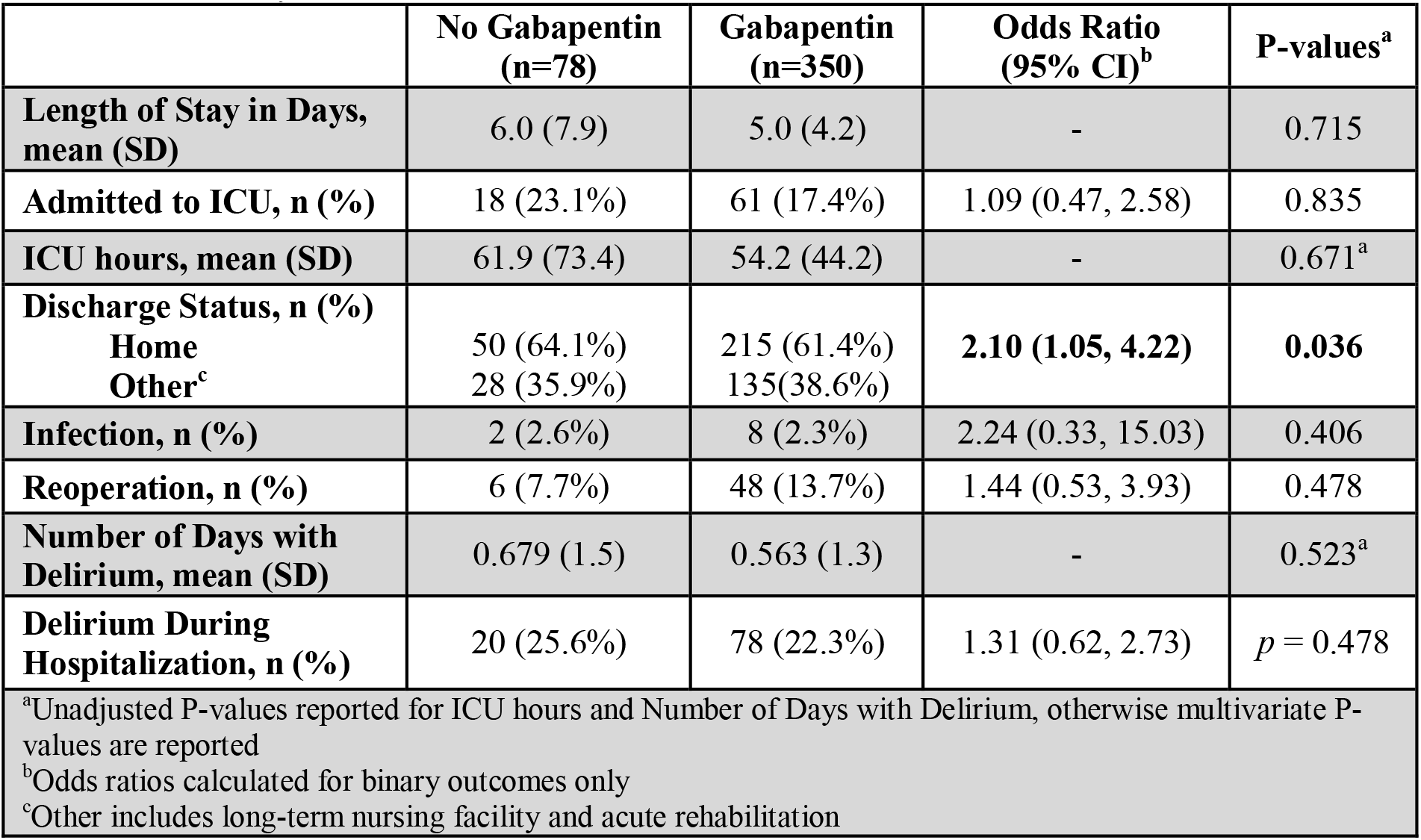
Secondary Outcome Measures.

|  | **No Gabapentin**<br>**(n=78)** | **Gabapentin**<br>**(n=350)** | **Odds Ratio**<br>**(95% CI)<sup>b</sup>** | **P-values<sup>a</sup>** |
| --- | --- | --- | --- | --- |
| **Length of Stay in Days,**<br>**mean (SD)** | 6.0 (7.9) | 5.0 (4.2) | - | 0.715 |
| **Admitted to ICU, n (%)** | 18 (23.1%) | 61 (17.4%) | 1.09 (0.47, 2.58) | 0.835 |
| **ICU hours, mean (SD)** | 61.9 (73.4) | 54.2 (44.2) | - | 0.671<sup>a</sup> |
| **Discharge Status, n (%)** |  |  |  |  |
| Home | 50 (64.1%) | 215 (61.4%) | **2.10 (1.05, 4.22)** | **0.036** |
| Other<sup>c</sup> | 28 (35.9%) | 135 (38.6%) |  |  |
| **Infection, n (%)** | 2 (2.6%) | 8 (2.3%) | 2.24 (0.33, 15.03) | 0.406 |
| **Reoperation, n (%)** | 6 (7.7%) | 48 (13.7%) | 1.44 (0.53, 3.93) | 0.478 |
| **Number of Days with**<br>**Delirium, mean (SD)** | 0.679 (1.5) | 0.563 (1.3) | - | 0.523<sup>a</sup> |
| **Delirium During**<br>**Hospitalization, n (%)** | 20 (25.6%) | 78 (22.3%) | 1.31 (0.62, 2.73) | *p* = 0.478 |
| <sup>a</sup>Unadjusted P-values reported for ICU hours and Number of Days with Delirium, otherwise multivariate P-values are reported |  |  |  |  |
| <sup>b</sup>Odds ratios calculated for binary outcomes only |  |  |  |  |
| <sup>c</sup>Other includes long-term nursing facility and acute rehabilitation |  |  |  |  |

## DISCUSSION

### Discussion

Verret *et al*. recently investigated the effect of postoperative gabapentin in 281 clinical trials from various surgical populations. Their study represents one of the largest meta-analyses on this subject to date and concluded the use of gabapentin did not cause a clinically significant reduction in pain or opioid use.^26^ However, gabapentin remains a popular postoperative analgesic with many spine surgeons. This may be explained by the fact that literature investigating the use of gabapentin in spine surgery patients has shown gabapentin administration reduces postoperative pain, opioid requirements, and opioid-related side effects.^12,29,30^ Given the conflicting findings in surgical literature, the purpose of the current study was to investigate the effect of perioperative gabapentin in a modern MMA protocol for spine surgery patients. Our analysis did not find perioperative gabapentin administration reduced postoperative pain or opioid use. However, the use of gabapentin did not seem to increase the rates of major cognitive side effects as measured by the incidence of delirium.

Compared to other surgical populations, the spine surgery population frequently experiences chronic pain and related comorbidities such as depression. A lack of generalizability between surgical populations could partially explain the conflicting results in surgical literature regarding the efficacy of gabapentin. Other explanations that have been offered include issues with methodology in early studies such as insufficient blinding.^31^ The difference in findings in our cohort compared to prior studies of spine surgery patients could also be explained by surgical complexity. Our cohort predominantly consisted of multi-level lumbar fusions while other studies examined patients who underwent less high-risk procedures such as single level discectomies.^30,32^ It may be the case that postoperative pain in certain cases exhibits an increased response to gabapentin. Future studies may consider stratifying their analysis by the type of surgery and indication.

The multivariate analysis of postoperative pain and opioid use also found that the use of postoperative muscle relaxants was associated with higher reported pain scores, which has not been previously described. It is possible that patients with higher pain were more likely to receive muscle relaxants postoperatively. However, this seems unlikely since muscle relaxants were administered as standard protocol and both study groups had similar demographic and surgical characteristics. Inclusion of other potential confounders in the multivariate analysis demonstrated both outpatient antidepressant and outpatient opioid use were associated with higher postoperative pain scores which is in agreement with prior literature.^33^

Although VAS pain scores can be biased owing to the subjective nature of the measurement, several studies have confirmed that it remains one of the best patient-reported outcome measures for pain.^34,35^ This study also considered several secondary outcomes which may be impacted by pain and opioid usage. We did not find a difference between groups in terms of length of stay, ICU admission rates or rates of delirium. Patients in the gabapentin group were significantly more likely to be discharged to an acute care facility or long-term rehabilitation compared to the no gabapentin group. A potential explanation for this finding is that physicians began increasing the use of physical therapy and rehabilitation as part of their postoperative management around the time that the MMA protocols were implemented in our hospital.

### Limitations

This study has several limitations. First, this was a retrospective study and thus subject to inherent limitations owing to its retrospective design. Particularly, there is the possibility of channeling bias since analgesia protocols may have been altered based on clinical circumstances during the perioperative period. This study was also unable to assess outcomes regarding opioid related adverse effects, such as constipation, ileus, and nausea due to this information being unavailable in medical records. Prior studies have shown that gabapentin decreases opioid related side effects such as constipation, ileus, nausea, and vomiting, so we cannot rule out the possibility that patients in this study may have benefited from gabapentin use through a reduction in adverse drug effects.^30^ Additionally, patients were discharged on different postoperative days, based on the determination of readiness by the clinical team. which could have impacted the data regarding their pain and opioid use. To control for this, alternative IPW models which limited mean pain and MED to the first three postoperative days were used. These models also did not demonstrate beneficial analgesic effects after gabapentin administration. Lastly, this cohort was restricted to patients over the age of 65 to avoid false positives during the health record query for degenerative spine disease and may not be generalizable to younger surgical populations.

## CONCLUSION

This retrospective cohort study of patients who underwent elective posterior lumbar fusion for degenerative spine disease did not find perioperative gabapentin administration to be beneficial in reducing postoperative pain or opioid use. The total length of stay, ICU admission rates, and rates of delirium were comparable between study groups. Patients who received gabapentin were more likely to be discharged to a rehabilitation or skilled care facility, although this may reflect increased utilization of these resources by providers. These findings provide additional support to recent evidence that gabapentin has minimal benefit in pain management during the acute postoperative period.

## Data Availability

All data produced in the present study are available upon reasonable request to the authors

## Acknowledgements

Thank you to Gregory Hopson and Linda Kleinmeyer for their assistance with the acquisition of the data from the electronic health records

## Notes

**Disclosures:** The authors have no conflicts of interest or funding sources to disclose

### Competing Interest Statement

The authors have declared no competing interest.

### Funding Statement

This study did not receive any funding

### Author Declarations

IRB of the University of Iowa waived ethical approval for this work.

